# Antihypertensive Medications and COVID-19 Diagnosis and Mortality: Population-based Case-Control Analysis in the United Kingdom

**DOI:** 10.1101/2020.09.25.20201731

**Authors:** Emma Rezel-Potts, Abdel Douiri, Phil J. Chowienczyk, Martin C. Gulliford

## Abstract

**Objectives:** To evaluate antihypertensive medications and COVID-19 diagnosis and mortality, accounting for healthcare seeking behaviour.

**Design:** A population-based case control study with additional cohort analysis.

**Setting:** Primary care patients from the UK Clinical Practice Research Datalink (CPRD).

**Participants:** 16 866 patients with COVID-19 events in the CPRD from 29^th^ January to June 25^th^ 2020 and 70 137 matched controls.

**Main outcome measures:** We explored associations between COVID-19 diagnosis and prescriptions for angiotensin converting enzyme inhibitors (ACEIs), angiotensin receptor blockers (ARBs), beta-blockers (B), calcium-channel blockers (C), thiazide diuretics (D) and other antihypertensive drugs (O). We evaluated all-cause mortality among COVID-19 cases. Analyses were adjusted for covariates and consultation frequency.

**Results:** In covariate adjusted analyses, ACEIs were associated with lower odds of COVID-19 diagnosis (0.82, 95% confidence interval 0.77 to 0.88) as were ARBs, 0.87 (0.80 to 0.95) with little attenuation from adjustment for consultation frequency. In fully adjusted analyses, C and D were also associated with lower odds of COVID-19. Increased odds of COVID-19 for B (1.19, 1.12 to 1.26), were attenuated after adjustment for consultation frequency (1.01, 0.95 to 1.08). In adjusted analyses, patients treated with ACEIs or ARBs had similar mortality to patients treated with classes B, C, D or O (1.00, 0.83 to 1.20) or patients receiving no antihypertensive therapy (0.99, 0.83 to 1.18).

**Conclusions:** Associations were sensitive to adjustment for confounding and healthcare seeking, but there was no evidence that antihypertensive therapy is associated with increased risk of COVID-19 diagnosis or mortality; most classes of antihypertensive therapy showed negative associations with COVID-19 diagnosis.

## INTRODUCTION

Coronavirus disease 2019 (COVID-19) caused by the novel severe acute respiratory syndrome coronavirus 2 (SARS-CoV-2) has become a global pandemic. The World Health Organization’s 29 June 2020 situation report documented 10 million cases and nearly 500 000 deaths of COVID-19 globally.^1^ The United Kingdom is among the countries with the highest COVID-19 incidence and death rates in the world, with figures from 14 September 2020 indicating that among those first testing positive for the virus, in excess of 41 637 died within 28 days.^2^

Biomedical scientific evidence suggests a role of the renin-angiotensin-aldosterone system (RAAS) in COVID-19. SARS-CoV-2 enters host cells via interaction with the angiotensin-converting enzyme 2 (ACE2) receptor which is part of the renin-angiotensin-aldosterone system (RAAS).^3-6^ Angiotensin-converting enzyme inhibitors (ACEIs) and angiotensin receptor blockers (ARBs) modulate the RAAS system and treatment with ACEIs and ARBs may enhance ACE2 activity thereby increasing SARS-CoV-2 susceptibility and COVID-19 severity.^7 8^ Conversely, increased ACE2 might have a protective effect by competitive inhibition of SARS-CoV-2 entry into respiratory epithelium or via negative regulation of the RAAS for anti-inflammatory, antioxidative and vasodilatory effects.^7 9^

Antihypertensive drugs represent the most frequently prescribed medicines in the UK, used by 15% of adults with long term conditions including diabetes, hypertension and cardiovascular diseases.^10^ These pre-existing conditions are frequent among those receiving healthcare for COVID-19^6 11^ and have also been associated with high COVID-19 case fatality rates,^12-14^ raising concerns about possible associations with AHT. There have been conflicting results from several observational studies exploring the relationship between AHT and COVID-19 susceptibility and severity since the start of the pandemic.^15 16^

A living systematic review, updated to 3^rd^ August 2020, included three studies with 8,766 COVID-19 patients and found no evidence of association between either ACEI or ARB treatment and positive COVID-19 test results.^15^ Two of the studies drew on patients attending hospital in the US,^17 18^ while the third study was population-based, including both primary care and hospital attendances in the Lombardy region in Italy.^19^ A large international study which was initially included in the review has since been retracted by the journal in which it was published. A retrospective cohort study using Danish national disease registries which found no evidence of association of prior treatment with ACEIs/ARBs with COVID-19 diagnosis, severity or death.^20^ In contrast, a systematic review and meta-analysis of 28,872 patients found that treatment with RAAS inhibitors (RAASi) was associated with lower risk of death or critical events.^16^ A primary care database study in the UK reported that prior use of ACEIs and ARBs were associated with lower risk of COVID-19 diagnosis, but there was no association with intensive care unit admission.^21^

Non-randomised studies to evaluate the association of RAASi drugs on COVID-19 susceptibility and severity face several methodological challenges. Firstly, there may be substantial confounding. Several previous studies have only presented ‘fully adjusted’ estimates, making it difficult to assess the possibility of residual confounding. Secondly, analysis of clinical data from electronic health records may be associated with misclassification of confounders, which might bias estimates in either positive or negative directions. Thirdly, and perhaps most importantly, cases of COVID-19 documented into electronic health records represent only a minority of cases occurring in the community during the first wave of the pandemic. If hypertension and COVID-19 are both associated with increased health care utilisation, this might introduce spurious associations between antihypertensive therapy and COVID-19 through collider bias.^22 23^ Selection pressures may have biased the sample toward those with greater symptom severity^24^ or who have comorbidities that make contact with health services more likely.

This study aimed to add to the evidence concerning antihypertensive therapy and COVID-19 diagnosis and mortality through analysis of a large population-based sample registered in UK primary care. We aimed to evaluate the association between AHT classes including ACEIs, ARBs, calcium-channel blockers, beta-blockers and thiazide diuretics and COVID-19 diagnosis and mortality. We aimed to explore the effects of covariate adjustment including patients’ consultation frequency in primary care.

## METHODS

### Overview

We undertook a case-control analysis of CPRD GOLD to estimate associations between antihypertensive treatments and COVID-19 susceptibility. We examined associations between antihypertensive treatments and COVID-19 mortality using date of death in the CPRD within 30 days as the outcome. Analyses were adjusted for covariates and consultation frequency.

### Case and control selection

The CPRD GOLD is among the world’s largest databases of anonymised electronic health records from primary care. It is estimated that 7% of UK general practices contribute to CPRD GOLD enabling the database to have extensive coverage and good sociodemographic and geographic representativeness of the UK population.^25^ The high quality of the CPRD GOLD data has been verified by several studies.^26^ The protocol was approved by the CPRD Independent Scientific Advisory Committee (ISAC protocol 20_081RA).

Patients recorded with COVID-19 were identified from the July 2020 release of CPRD GOLD. COVID-19 events were identified from Read codes recorded into patients’ clinical, referral and test records: ‘Suspected disease caused by 2019-nCoV (novel coronavirus)’, 39%; ‘Suspected coronavirus infection’, 19%; ‘Telephone consultation for suspected 2019-nCoV (novel coronavirus)’, 16%; ‘2019-nCoV (novel coronavirus) detected’, 12%; ‘Disease caused by 2019-nCoV (novel coronavirus)’, 7%; ‘Coronavirus infection’, 5%; ‘Coronavirus nucleic acid detection’, 3%; ‘[X] Coronavirus infection, unspecified’ and ‘Coronavirus as cause of disease classified to other chapters’, less than 1%. We excluded the small number of COVID-19 diagnosis and mortality events dated on or before 29^th^ January 2020, the official date of the UK’s first confirmed COVID-19 case.

COVID-19 cases were compared with a sample of matched control patients who did not have a COVID-19 diagnosis. Up to five control patients for each case were randomly sampled from the entire registered population of CPRD GOLD, matching on age, gender, index date and general practice.

### Exposures

The exposure was defined as prescription of antihypertensive treatments (AHT) in the six months before the index date of the following classes: ACEIs, ARBs, beta blockers (B), calcium channel blockers (C), thiazide diuretics (D) and other antihypertensive drugs (O). Each class of AHT was coded as either prescribed or not prescribed.

### Covariates

Covariates were defined using data recorded in the five-year period before date of diagnosis. These included smoking status (non-smoker, current smoker, ex-smoker), body mass index (BMI, underweight, normal weight, overweight, obese, and missing), systolic blood pressure (SBP) and diastolic blood pressure (DBP) in categories of 10 mm Hg. Ethnicity was classified as ‘white’, ‘black’, ‘Asian’, ‘mixed’, ‘other’ and ‘not known’. We used Clegg’s electronic Frailty Index (eFI) to evaluate frailty into categories of non-frail, mild, moderate and severe frailty ^27^. We also evaluated the 15 comorbidities of the Charlson comorbidity index as present or absent, following the recommendations of Khan et al.^28 29^ We evaluated healthcare seeking behaviour by calculating the rate of events in each patient’s clinical record file, including consultations and other contacts with the practice, in the year preceding the index date. This rate per patient year was entered as a continuous variable.

### Analysis

A conditional logistic regression model was employed for the case control analysis. Unadjusted odds ratios were compared with covariate adjusted odds ratios including ethnicity, BMI, blood pressure, smoking status, frailty level, comorbidities and treatment with each class of AHT. Finally, additional adjustment was made for consultation frequency. As a sensitivity analysis we repeated the analysis only including patients with confirmed COVID-19 diagnoses, excluding patients recorded to have ‘suspected’ COVID-19. We also explored evidence of an interaction between age group and each class of AHT, conducting a sub-group analysis to examine age as an effect modifier.

A Cox proportional hazards model was employed to examine the association of AHT with patient survival. Survival time in days from the date of first COVID-19 diagnosis to date of death or end of record was the outcome. Patients who were still alive at the end of the study period were censored. Covariates were month of COVID-19 diagnosis, gender, age (zero to four, five to nine and 10 to 14 and then 10 years age groups up to 85 years and over), and smoking status, BMI, blood pressure, ethnic group, eFI category comorbidities and treatment with each class of AHT. A secondary analysis also adjusted for region of practice. We did not adjust for consultation frequency because all patients in this analysis had already accessed care from their general practice. We also evaluated ACEIs and ARBs combined, as renin-angiotensin-aldosterone system inhibiting drugs (RAASi), making comparisons with other classes of AHT drugs (BCDO) or no antihypertensive treatment. The proportional hazards assumption was checked graphically and with a test for proportional hazards. All data were analysed in R, version 4.0.2.

## RESULTS

### Case-control analysis: COVID-19 diagnosis

There were 16 866 COVID-19 cases identified in CPRD GOLD from 29 January to 20 June 2020 (Supplementary Figure 1), which were compared to 70 137 matched controls. COVID-19 cases had consulted their general practice more frequently over the preceding year than controls, with a mean rate of clinical record events in the preceding year of 28.0 per person year, compared to 15.4 per person year among controls.

Table 1 and Supplementary Table 1 show descriptive data for COVID-19 cases and controls as well as unadjusted and covariate adjusted odds ratios and 95% confidence intervals.

**Table 1:**
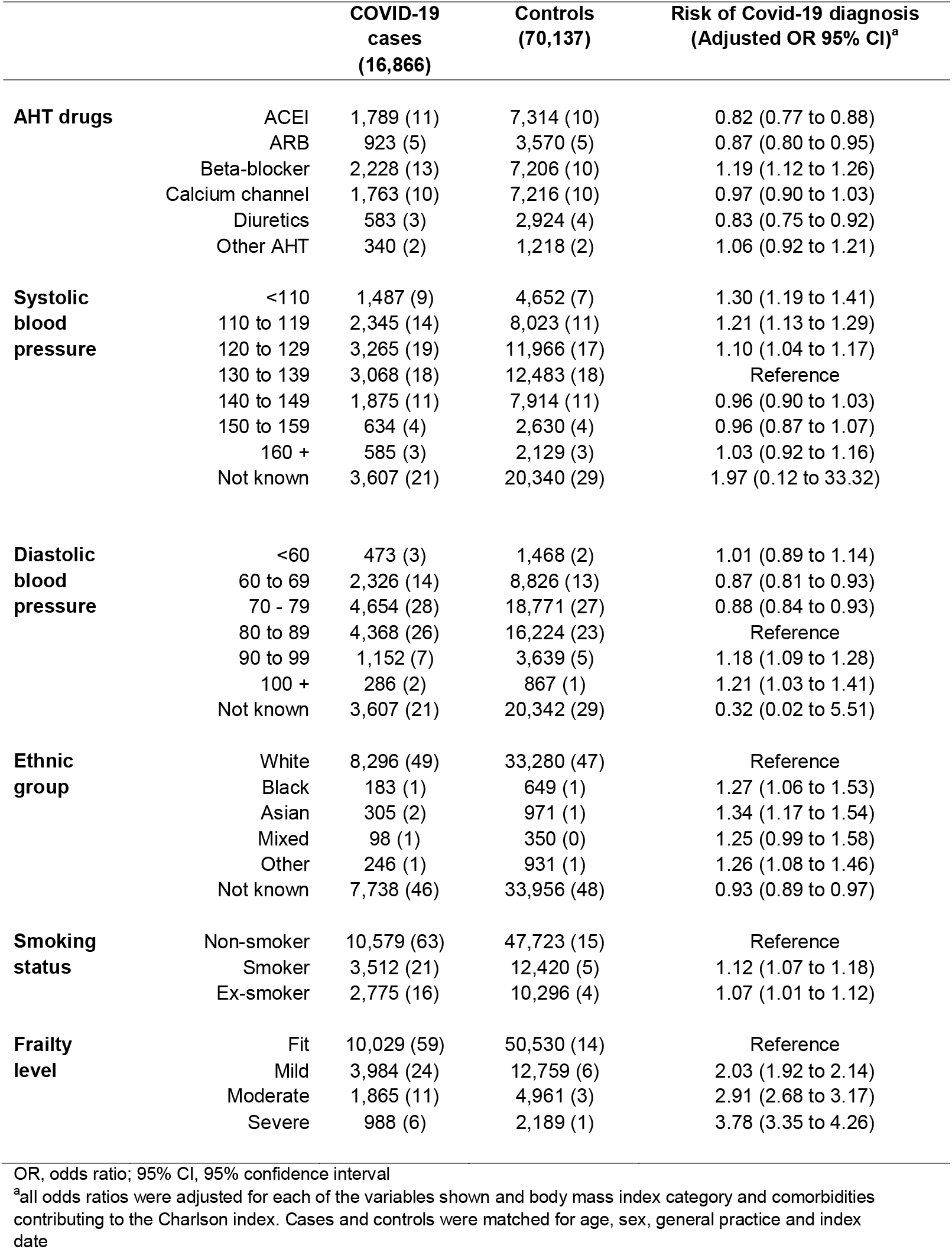
Variables associated with Covid-19 diagnosis. Figures are frequencies (column percent) except where indicated.

|  |  | **COVID-19 cases**<br>**(16,866)** | **Controls**<br>**(70,137)** | **Risk of Covid-19 diagnosis**<br>**(Adjusted OR 95% CI)<sup>a</sup>** |
| --- | --- | --- | --- | --- |
| **AHT drugs** | ACEI | 1,789 (11) | 7,314 (10) | 0.82 (0.77 to 0.88) |
|  | ARB | 923 (5) | 3,570 (5) | 0.87 (0.80 to 0.95) |
|  | Beta-blocker | 2,228 (13) | 7,206 (10) | 1.19 (1.12 to 1.26) |
|  | Calcium channel | 1,763 (10) | 7,216 (10) | 0.97 (0.90 to 1.03) |
|  | Diuretics | 583 (3) | 2,924 (4) | 0.83 (0.75 to 0.92) |
|  | Other AHT | 340 (2) | 1,218 (2) | 1.06 (0.92 to 1.21) |
| **Systolic blood pressure** | <110 | 1,487 (9) | 4,652 (7) | 1.30 (1.19 to 1.41) |
|  | 110 to 119 | 2,345 (14) | 8,023 (11) | 1.21 (1.13 to 1.29) |
|  | 120 to 129 | 3,265 (19) | 11,966 (17) | 1.10 (1.04 to 1.17) |
|  | 130 to 139 | 3,068 (18) | 12,483 (18) | Reference |
|  | 140 to 149 | 1,875 (11) | 7,914 (11) | 0.96 (0.90 to 1.03) |
|  | 150 to 159 | 634 (4) | 2,630 (4) | 0.96 (0.87 to 1.07) |
|  | 160 + | 585 (3) | 2,129 (3) | 1.03 (0.92 to 1.16) |
|  | Not known | 3,607 (21) | 20,340 (29) | 1.97 (0.12 to 33.32) |
| **Diastolic blood pressure** | <60 | 473 (3) | 1,468 (2) | 1.01 (0.89 to 1.14) |
|  | 60 to 69 | 2,326 (14) | 8,826 (13) | 0.87 (0.81 to 0.93) |
|  | 70 - 79 | 4,654 (28) | 18,771 (27) | 0.88 (0.84 to 0.93) |
|  | 80 to 89 | 4,368 (26) | 16,224 (23) | Reference |
|  | 90 to 99 | 1,152 (7) | 3,639 (5) | 1.18 (1.09 to 1.28) |
|  | 100 + | 286 (2) | 867 (1) | 1.21 (1.03 to 1.41) |
|  | Not known | 3,607 (21) | 20,342 (29) | 0.32 (0.02 to 5.51) |
| **Ethnic group** | White | 8,296 (49) | 33,280 (47) | Reference |
|  | Black | 183 (1) | 649 (1) | 1.27 (1.06 to 1.53) |
|  | Asian | 305 (2) | 971 (1) | 1.34 (1.17 to 1.54) |
|  | Mixed | 98 (1) | 350 (0) | 1.25 (0.99 to 1.58) |
|  | Other | 246 (1) | 931 (1) | 1.26 (1.08 to 1.46) |
|  | Not known | 7,738 (46) | 33,956 (48) | 0.93 (0.89 to 0.97) |
| **Smoking status** | Non-smoker | 10,579 (63) | 47,723 (15) | Reference |
|  | Smoker | 3,512 (21) | 12,420 (5) | 1.12 (1.07 to 1.18) |
|  | Ex-smoker | 2,775 (16) | 10,296 (4) | 1.07 (1.01 to 1.12) |
| **Frailty level** | Fit | 10,029 (59) | 50,530 (14) | Reference |
|  | Mild | 3,984 (24) | 12,759 (6) | 2.03 (1.92 to 2.14) |
|  | Moderate | 1,865 (11) | 4,961 (3) | 2.91 (2.68 to 3.17) |
|  | Severe | 988 (6) | 2,189 (1) | 3.78 (3.35 to 4.26) |

OR, odds ratio; 95% CI, 95% confidence interval
<sup>a</sup>all odds ratios were adjusted for each of the variables shown and body mass index category and comorbidities contributing to the Charlson index. Cases and controls were matched for age, sex, general practice and index date

COVID-19 diagnosis was associated with current smoking, underweight and obese BMI, frailty and black and minority ethnicity. Both low and high systolic or diastolic blood pressure values were associated with increased odds of COVID-19 diagnosis when compared with intermediate blood pressure values. Comorbidities associated with COVID-19 diagnosis in covariate adjusted analyses included chronic pulmonary disease, metastatic tumour, congestive heart disease, cerebrovascular disease and dementia. Diabetes and renal disease were associated with COVID-19 diagnosis in unadjusted but not covariate adjusted analyses.

There were 4 852 (29%) COVID-19 cases prescribed antihypertensive treatment (AHT) in the year preceding diagnosis, compared to 17 978 (26%) of controls. Positive associations of ACEI and ARB with COVID-19 diagnosis in unadjusted analyses (Supplementary Table 1) were attenuated in the covariate adjusted analysis (Table 1), with both ACEIs and ARBs being negatively associated with COVID-19 diagnosis (ACEI: odds ratio 0.82, 95% CI 0.77 to 0.88; ARB: 0.87, 0.80 to 0.95). Further adjustment for consultation frequency had little effect on estimated associations (ACEI: 0.80, 0.75 to 0.86; ARB: 0.84, 0.77 to 0.92) (Figure 1). In the fully adjusted model, calcium channel blockers and thiazide diuretics were also associated with lower odds of COVID-19 diagnosis (Figure 1). In covariate adjusted analyses, beta-blocker treatment was positively associated with COVID-19 diagnosis (1.19, 1.12 – 1.26) but this was attenuated after further adjustment for consultation frequency (1.01, 0.95 to 1.08) (Figure 1). When the sample was restricted to only those cases with confirmed COVID-19 diagnoses and their matched controls, there were 4 233 cases matched to 17 700 controls, the pattern of association was unaltered though estimates were necessarily less precise (Supplementary Figure 3).

**Figure 1:**
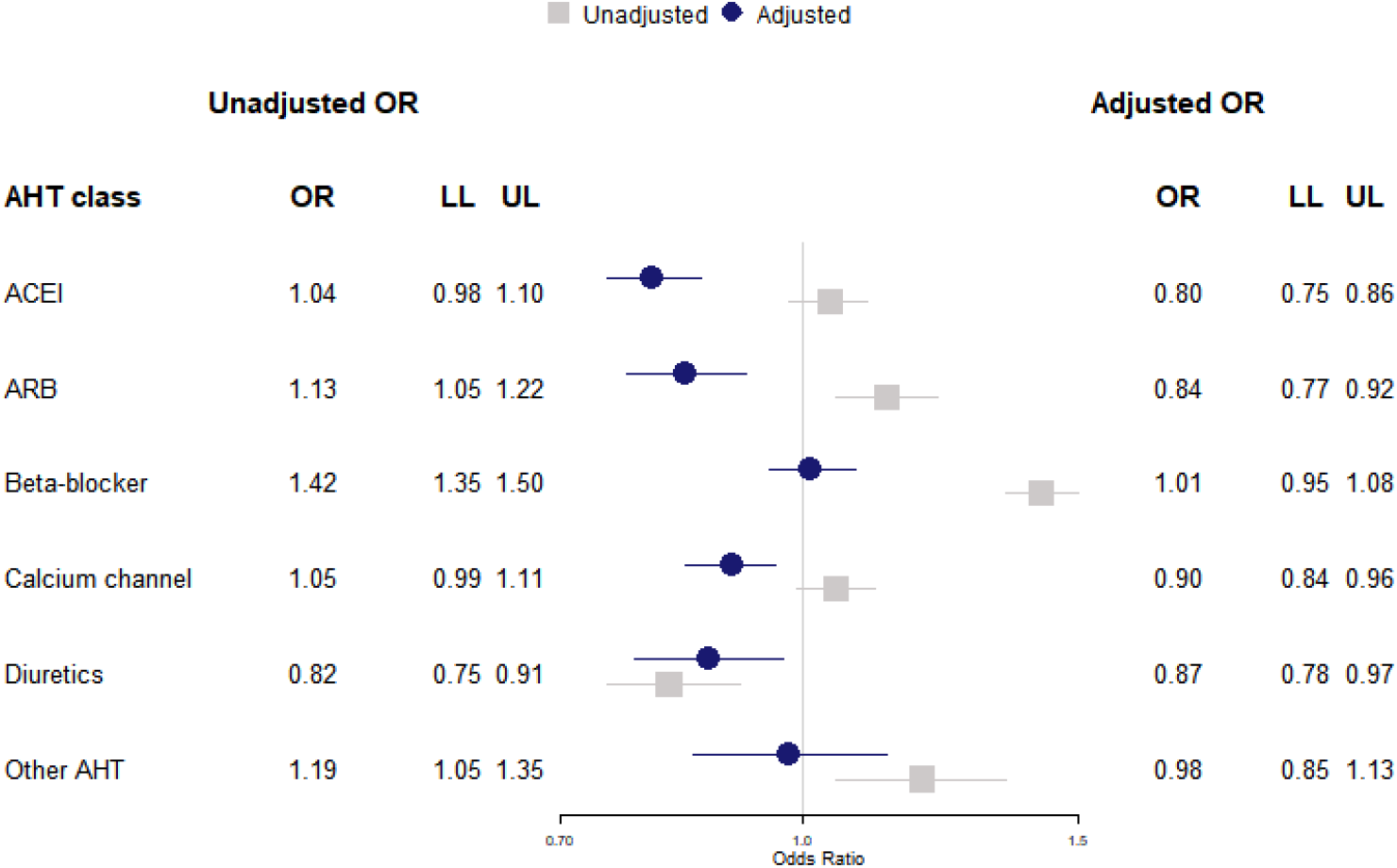
Case-control analysis for COVID-19 diagnosis showing unadjusted odds ratios (grey) and model adjusted for ethnicity, body mass index, blood pressure, smoking status, frailty level, comorbidities and the rate of events in each patient’s clinical record in the year preceding the index date (blue). OR, odds ratio; LL, lower limit 95% confidence interval; UL, upper limit 95% confidence interval; ACEI, angiotensin-converting enzyme inhibitors; ARB, angiotensin receptor blockers

Adding an interaction term for age within logistic regression models for each class of AHT improved the model goodness of fit, therefore we conducted a subgroup analysis for cases and controls by AHT treatment and age (Supplementary Table 3). Whilst lower age categories had very few observations, AHT therapy was positively associated with higher odds of COVID-19 diagnosis across all classes ABCDO within the mid-range age categories, such as those aged 45-54 years on ACEI (1.47, 1.27 to 1.69). These associations were attenuated after adjusting for all covariates including consultation frequency.

### Primary cohort analysis: COVID-19 mortality

Among the 16 866 COVID-19 cases, 921 (5%) died within 30 days. Patients treated with antihypertensive drugs were more highly represented among deceased patients than the overall sample and this was true for each class of AHT drugs (Table 2 and Supplementary Table 2). Table 2 and Supplementary Table 2 provide estimates for covariates which show that male gender, older age, black and ethnic minority status, diabetes, metastatic tumour, dementia and mild liver disease were associated with greater mortality. In covariate adjusted analyses, there was no evidence that any of the classes of AHT drugs might be associated with mortality after COVID-19 diagnosis (Table 2). Additional adjustment for practice region did not influence associations (Figure 2). There was no evidence that treatment with RAASi drugs, including ACEI and ARB drugs, might be associated with higher mortality than other classes of AHT drugs (BCDO) with hazard ratio 1.00 (0.83 to 1.20). Similarly there was no evidence that treatment with RAASi drugs was associated with greater risk than no antihypertensive treatment (0.99, 0.83 to 1.18), nor was treatment with BCD and O classes associated with higher mortality than no AHT treatment (0.99, 0.84 to 1.18).

**Table 2:** Variables associated with Covid-19 diagnosis. Figures are frequencies (column percent) except where indicated.

|  |  | **COVID-19**<br>**cases**<br>**(16,866)** | **Death in 30**<br>**days**<br>**921 (5)** | **Adjusted HR**<br>**(95% CI)** |
| --- | --- | --- | --- | --- |
| **AHT drugs** | ACEI | 1,789 (11) | 182 (20) | 1.08 (0.90 to 1.30) |
|  | ARB | 923 (5) | 72 (8) | 0.84 (0.65 to 1.09) |
|  | Beta-blocker | 2,228 (13) | 263 (29) | 1.13 (0.97 to 1.33) |
|  | Calcium channel | 1,763 (10) | 149 (16) | 0.87 (0.72 to 1.04) |
|  | Diuretics | 583 (3) | 47 (5) | 1.00 (0.74 to 1.36) |
|  | Other AHT | 340 (2) | 30 (3) | 0.85 (0.59 to 1.24) |
| **Gender** | Male | 6,796 (40) | 465 (50) | 1.51 (1.32 to 1.74) |
|  | Female | 10,070 (60) | 456 (50) | Reference |
| **Age group**<br>**(years)** | 0-4 | 697 (4) | 0 (0) | 0.00 (0.00 to Inf.) |
|  | 5-14 | 641 (4) | 0 (0) | 0.00 (0.00 to Inf.) |
|  | 15-24 | 889 (5) | 2 (0) | 0.10 (0.02 to 0.41) |
|  | 25-34 | 1,959 (12) | 0 (0) | 0.00 (0.00 to Inf.) |
|  | 35-44 | 2,259 (13) | 6 (1) | 0.12 (0.05 to 0.27) |
|  | 45-54 | 2,663 (16) | 13 (1) | 0.23 (0.12 to 0.42) |
|  | 55-64 | 2,747 (16) | 56 (6) | Reference |
|  | 65-74 | 1,701 (10) | 126 (14) | 3.52 (2.56 to 4.85) |
|  | 75-84 | 1,662 (10) | 310 (34) | 8.18 (6.02 to 11.10) |
|  | 85+ | 1,648 (10) | 408 (44) | 10.74 (7.83 to 14.72) |
| **Systolic blood**<br>**pressure** | <110 | 1,487 (9) | 109 (12) | 1.30 (0.99 to 1.70) |
|  | 110 - 119 | 2,345 (14) | 128 (14) | 1.23 (0.97 to 1.56) |
|  | 120 to 129 | 3,265 (19) | 171 (19) | 0.98 (0.79 to 1.21) |
|  | 130 to 139 | 3,068 (18) | 181 (20) | Reference |
|  | 140 to 149 | 1,875 (11) | 130 (14) | 0.93 (0.74 to 1.17) |
|  | 150 to 159 | 634 (4) | 45 (5) | 0.96 (0.69 to 1.34) |
|  | 160 + | 585 (3) | 46 (5) | 0.76 (0.54 to 1.08) |
|  | Not known | 3,607 (21) | 111 (12) | 5.78 (0.00 to ne) |
| **Diastolic blood**<br>**pressure** | <60 | 473 (3) | 83 (9) | 1.42 (1.05 to 1.91) |
|  | 60 to 69 | 2,326 (14) | 194 (21) | 1.05 (0.84 to 1.31) |
|  | 70 - 79 | 4,654 (28) | 276 (30) | 1.03 (0.85 to 1.24) |
|  | 80 to 89 | 4,368 (26) | 191 (21) | Reference |
|  | 90 to 99 | 1,152 (7) | 50 (5) | 1.36 (0.99 to 1.87) |
|  | 100 + | 286 (2) | 16 (2) | 1.12 (0.66 to 1.93) |
|  | Not known | 3,607 (21) | 111 (12) | 0.29 (0.00 to ne) |
| **Ethnic group** | White | 8,296 (49) | 396 (43) | Reference |
|  | Black | 183 (1) | 9 (1) | 2.60 (1.33 to 5.08) |
|  | Asian | 305 (2) | 9 (1) | 2.15 (1.10 to 4.19) |
|  | Mixed | 98 (1) | 1 (0) | 0.70 (0.10 to 5.05) |
|  | Other | 246 (1) | 5 (1) | 1.04 (0.43 to 2.52) |
|  | Not known | 7,738 (46) | 501 (54) | 1.05 (0.92 to 1.20) |
| **Smoking status** | Non-smoker | 10,579 (63) | 550 (60) | Reference |
|  | Smoker | 3,512 (21) | 304 (33) | 1.12 (0.96 to 1.30) |
|  | Ex-smoker | 2,775 (16) | 67 (7) | 0.81 (0.62 to 1.05) |
| **Frailty level** | Fit | 10,029 (59) | 182 (20) | Reference |
|  | Mild | 3,984 (24) | 265 (29) | 0.91 (0.74 to 1.12) |
|  | Moderate | 1,865 (11) | 283 (31) | 1.15 (0.91 to 1.45) |
|  | Severe | 988 (6) | 191 (21) | 1.16 (0.88 to 1.52) |

HR, hazard ratio; 95% CI, 95% confidence interval
<sup>a</sup>all odds ratios were adjusted for each of the variables shown and body mass index category and comorbidities contributing to the Charlson index, as well as age group, gender, general practice and month of diagnosis

**Figure 2.**
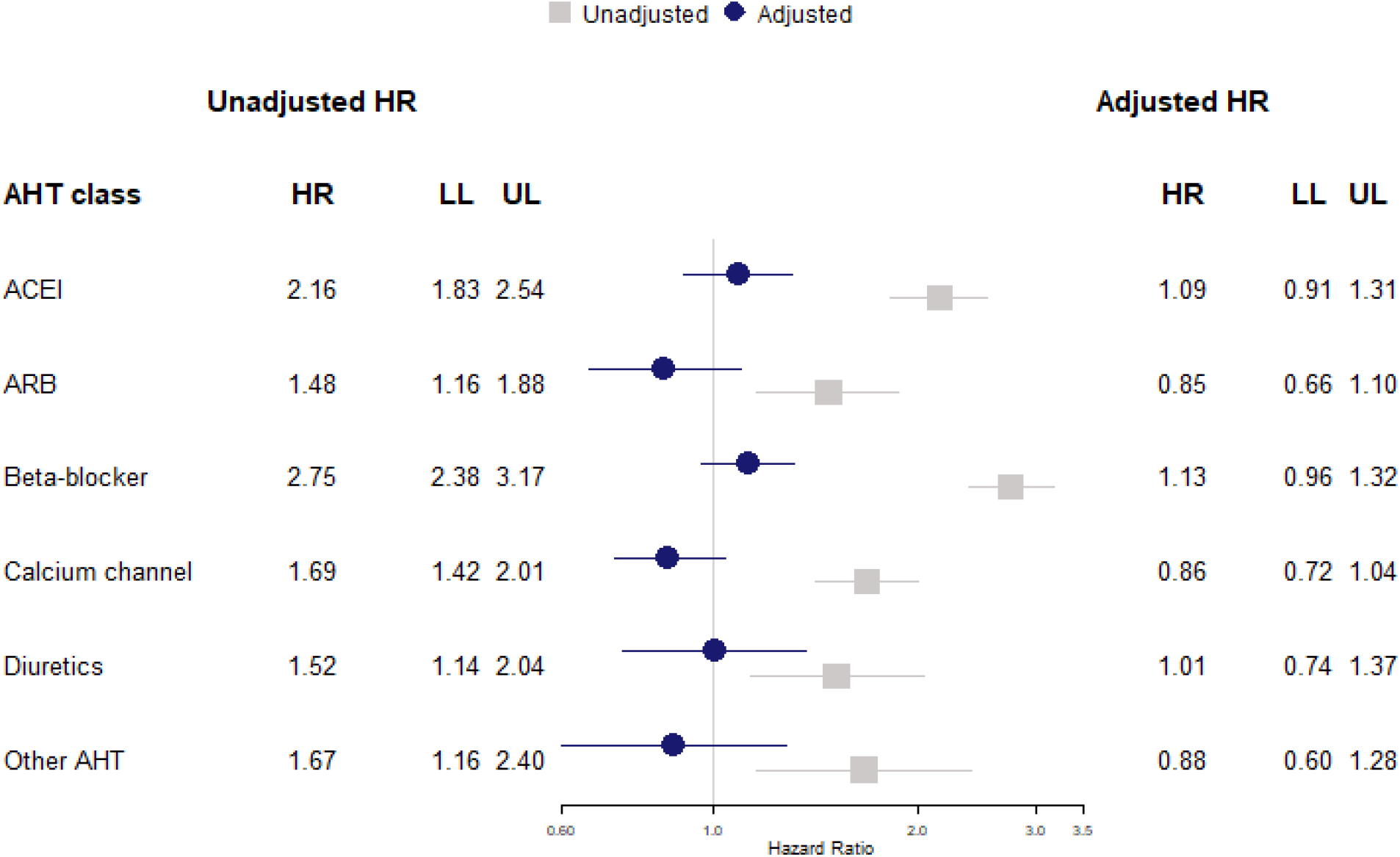
Cohort analysis for 30-day mortality following COVID-19 diagnosis showing unadjusted hazard ratios (grey) and model adjusted for body mass index, blood pressure, smoking status, frailty level, comorbidities and practice region (blue). HR, hazard ratio; LL, lower limit 95% confidence interval; UL, upper limit 95% confidence interval; AHT, antihypertensive treatment; ACEI, angiotensin-converting enzyme inhibitors; ARB, angiotensin receptor blockers

**Figure 3.**
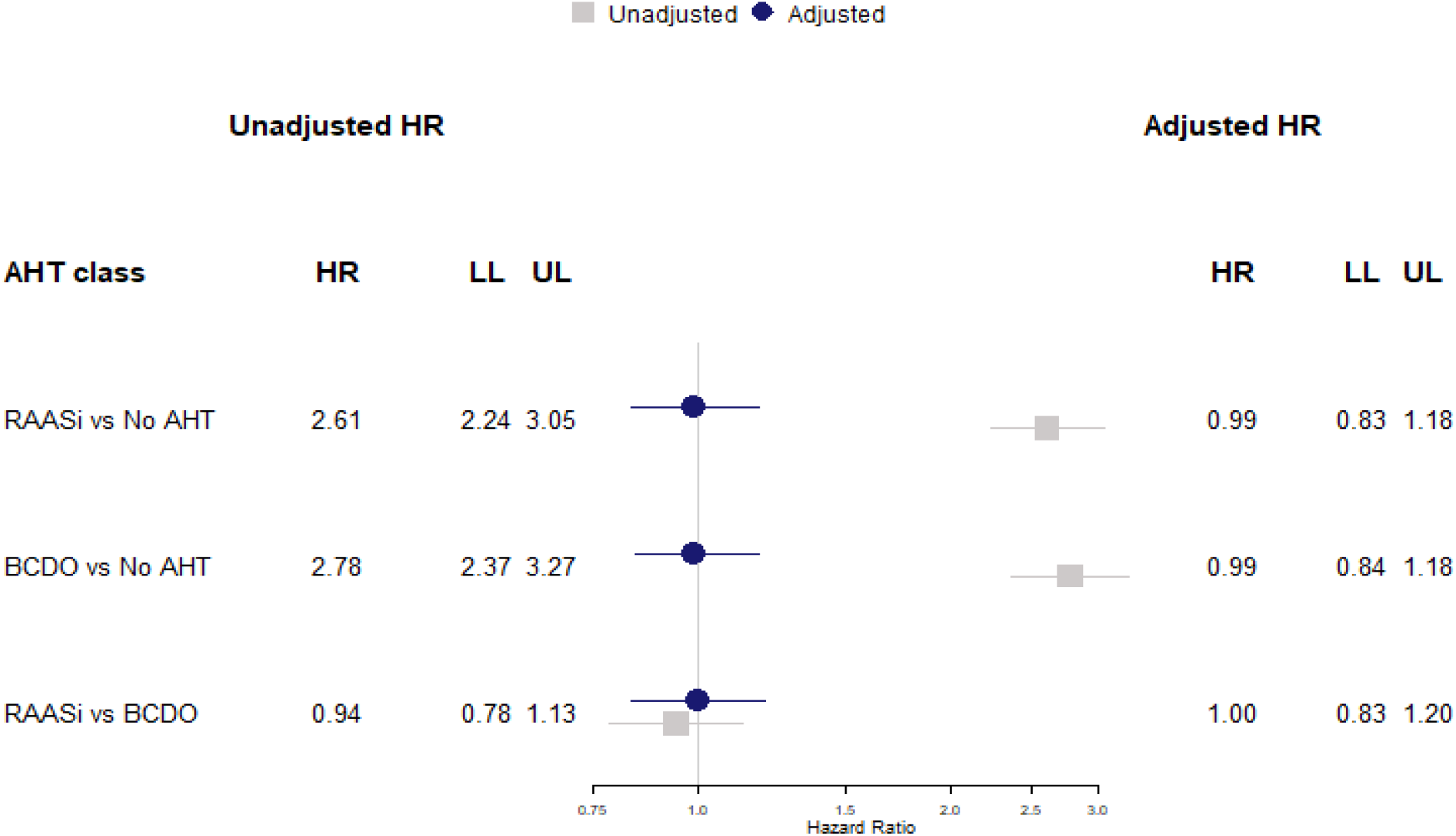
Cohort analysis for 30-day mortality following COVID-19 diagnosis showing unadjusted hazard ratios (grey) and model adjusted for body mass index, blood pressure, smoking status, frailty level, comorbidities (blue) comparing i) RAASi to no AHT ii) BCDO AHT (non-RAASi) to no AHT iii) RAASi to BCDO HR, hazard ratio; LL, lower limit 95% confidence interval; UL, upper limit 95% confidence interval; AHT, antihypertensive treatment; RAASi, renin-angiotensin-aldosterone system inhibitors; BCDO, beta-blocker, calcium-channel, diuretics, other

## DISCUSSION

### Statement of principle findings

This large population-based study included patients diagnosed with suspected or confirmed COVID-19 in UK primary care. During the first wave of the pandemic, there was limited testing capacity and most suspected cases remained unconfirmed; however, restricting our analysis to confirmed cases yielded similar results. After adjusting for covariates that characterise case-mix, we found no evidence that treatment with ACEIs, ARBs, calcium channel blockers, thiazide diuretics or other antihypertensive drugs might be associated with greater risk of COVID-19 diagnosis. There was evidence that beta-blocker treatment was associated with greater odds of COVID-19 diagnosis but additional adjustment for patients’ underlying consultation frequency removed this association, which might be attributable to collider bias. After adjusting for covariates, including blood pressure, and consultation frequency, ACEIs, ARBs, calcium channel blockers and thiazide diuretics were associated with lower odds of COVID-19 diagnosis. This might suggest that at a given level of blood pressure, patients treated with antihypertensive drugs may be at lower risk, but this pattern of association does not appear to be specific to any class of antihypertensive drug.

Associations of AHT with COVID-19 diagnosis may be modified by age, but the effects of adjusting for covariates, including comorbidities, suggest this attributable to confounding. In this cohort analysis of patients diagnosed in UK primary care with suspected or confirmed COVID-19, recorded mortality was considerably higher than the overall reported infection fatality ratio for this condition but there was no evidence that any class of antihypertensive drug might be associated with increased mortality risk, and the adjusted hazard for drugs acting on the renin-angiotensin-aldosterone system was similar to that for patients treated with other classes of AHT drugs or patients not receiving AHT therapy.

### Strengths and weaknesses of the study

This study drew on a large, longitudinal population-based data resource that enabled us to conduct a matched case-control analysis of risk factors for clinical COVID-19 diagnosis, as well as a cohort study of risk factors for COVID-19 mortality. As noted above, most cases were diagnosed clinically as suspected cases because of the limited capacity for testing in the early stage of the pandemic. General practice systems capture comprehensive data for all prescriptions issued by the general practice, so we can be confident that this exposure was accurately recorded. However, prescription utilisation may not be universal and non-adherence to prescribed medicines may be widespread. Data for covariates was not always completely recorded. Data on ethnicity were missing for almost half the sample as reported by others;^25^ an important limitation given the disproportionate impact COVID-19 has had on ethnic minority populations in the US and UK.^14 30 31^ COVID-19 susceptibility and severity have also been associated with measures of deprivation.^14^ Matching on practice may have accounted for differences in area-level deprivation to an extent, but deprivation based on participants’ home postcodes or individual-level deprivation measures might improve precision. It is known that observational studies on COVID-19 outcomes may be susceptible to collider bias.^22 23^ This can lead to spurious associations if both antihypertensive therapy and COVID-19 are associated with greater likelihood of general practice consultations.

Selection pressures may have biased the sample toward those with increased symptom severity^24^ or who are otherwise more likely to recognise COVID-19 symptoms and make contact with health services. We accounted for the effects of health-seeking behaviour by introducing into the analysis the rate of events in each patient’s clinical record in the year preceding the index date. There were substantial differences in consultation frequency between cases and controls and adjusting for this metric nullified an apparent association between beta-blocker use and COVID-19 diagnosis. However, randomisation allocation will be preferred to provide a higher level of evidence.

### Strengths and weaknesses in relation to other studies

A substantial number of observational studies have previously evaluated whether drugs acting on the renin-angiotensin system, including ACEIs and ARBs, might be associated with susceptibility, severity and mortality from COVID-19, but few studies have considered other classes of antihypertensive medications. Several systematic reviews have summarised the main findings, which generally find no evidence for worse COVID-19 outcomes in patients treated with these medications.^32-35^ Some studies suggest possible benefits from ACEI treatment.^21^ However, systematic reviews also highlight limitations of the evidence presented to date.^35^ A high proportion of studies has been based on data from patients admitted to hospital with COVID-19, sometimes with small samples. Many studies failed to include adequate adjustment for confounding. Few, if any, studies have directly addressed the question of collider bias, which distort associations when data are gathered from patients conditional upon their attendance for health care. Recently, COVID-19 has been evaluated in an ongoing randomised controlled trial of ramipril treatment.^36^ The analysis found no evidence for an effect of ramipril on COVID-19 incidence or severity but the analysis comprised 102 patients with 11 cases of COVID-19.^36^ Further trials are ongoing.^15^

## Conclusions

Drawing on data for a large population-based sample and using rigorous analytical methods, this study adds to the evidence that antihypertensive therapy may be safely continued during the SARS-CoV-2 pandemic. While previous studies have largely evaluated drugs acting on the renin-angiotensin system, the study found no evidence that any class of antihypertensive therapy might be associated with greater risk of COVID-19 diagnosis or mortality. There was evidence that, after adjusting for covariates including blood pressure, several classes of antihypertensive therapy might be associated with lower risk of a clinical COVID-19 diagnosis, but this pattern of association was not apparent for COVID-19 mortality. While this might be interpreted as evidence of a protective effect, in this observational study it is not possible to exclude the possibility that this pattern of association may be caused by bias.

## Data Availability

Requests for access to data from the study should be addressed to. All proposals requesting data access will need to specify planned uses with approval of the study team and CPRD before data release.

## Data sources

The study is based in part on data from the Clinical Practice Research Datalink obtained under license from the UK Medicines and Healthcare products Regulatory Agency. However, the interpretation and conclusions contained in this report are those of the authors alone.

## Author Contributions

ERP designed the study with advice from MG, AD and PC; ERP analysed the data and drafted the paper; MG, AD and PC reviewed the analysis and contributed to drafts of the paper. All authors approved the final draft. The corresponding author attests that all listed authors meet authorship criteria and that no others meeting the criteria have been omitted.

## REFERENCES

1. World Health Organization. Coronavirus Disease (COVID-19) Situation Report 161, 2020. https://www.who.int/docs/default-source/coronaviruse/situation-reports/20200629-covid-19-sitrep-161.pdf?sfvrsn=74fde64e_2 (accessed 25 September 2020).

2. Department of Health and Social Care, Public Health England. Coronavirus (COVID-19) in the UK, Deaths in the United Kingdom, 2020. https://coronavirus.data.gov.uk/deaths (accessed 14 September 2020).

3. Lu R, Zhao X, Li J, et al. Genomic characterisation and epidemiology of 2019 novel coronavirus: implications for virus origins and receptor binding. Lancet 2020;395(10224):565-74. doi:https://doi.org/10.1016/S0140-6736(20)30251-8.

4. Hoffmann M, Kleine-Weber H, Schroeder S, et al. SARS-CoV-2 Cell Entry Depends on ACE2 and TMPRSS2 and Is Blocked by a Clinically Proven Protease Inhibitor. Cell 2020;181(2):271-80.e8. doi:https://doi.org/10.1016/j.cell.2020.02.052

5. Wan Y, Shang J, Graham R, et al. Receptor Recognition by the Novel Coronavirus from Wuhan: an Analysis Based on Decade-Long Structural Studies of SARS Coronavirus. J. Virol 2020;94(7):e00127– 20. doi:10.1128/JVI.00127-20

6. Li B, Yang J, Zhao F, et al. Prevalence and impact of cardiovascular metabolic diseases on COVID-19 in China. Clin Res Cardiol 2020;109(5):531–38. doi:10.1007/s00392-020-01626-9

7. Bavishi C, Maddox TM, Messerli FH. Coronavirus Disease 2019 (COVID-19) Infection and Renin Angiotensin System Blockers. JAMA Cardiol 2020 doi:10.1001/jamacardio.2020.1282

8. Liu Y, Yang Y, Zhang C, et al. Clinical and biochemical indexes from 2019-nCoV infected patients linked to viral loads and lung injury. Sci. China Life Sci 2020;63(3):364–74. doi:10.1007/s11427-020-1643-8

9. Batlle D, Wysocki J, Satchell K. Soluble angiotensin-converting enzyme 2: a potential approach for coronavirus infection therapy? Clin. Sci 2020;134(5):543–45. doi:10.1042/cs20200163 [published Online First: 2020/03/14]

10. NHS Digital. Health Survey for England 2016 Prescribed medicines, 2017. https://files.digital.nhs.uk/pdf/3/c/hse2016-pres-med.pdf (accessed 28 August 2020).

11. Yang J, Zheng Y, Gou X, et al. Prevalence of comorbidities and its effects in patients infected with SARS-CoV-2: a systematic review and meta-analysis. Int. J. Infect. Dis 2020;94:91-95. doi:https://doi.org/10.1016/j.ijid.2020.03.017

12. Wu Z, McGoogan JM. Characteristics of and Important Lessons From the Coronavirus Disease 2019 (COVID-19) Outbreak in China: Summary of a Report of 72⍰14 Cases From the Chinese Center for Disease Control and Prevention. JAMA 2020;323(13):1239–42. doi:10.1001/jama.2020.2648

13. Team CC-R. Preliminary Estimates of the Prevalence of Selected Underlying Health Conditions Among Patients with Coronavirus Disease 2019 - United States, February 12-March 28, 2020. MMWR Morb Mortal Wkly Rep 2020;69(13):382–86. doi:10.15585/mmwr.mm6913e2

14. Williamson EJ, Walker AJ, Bhaskaran K, et al. Factors associated with COVID-19-related death using OpenSAFELY. Nature 2020;584(7821):430–36. doi:10.1038/s41586-020-2521-4

15. Mackey K, King VJ, Gurley S, et al. Risks and Impact of Angiotensin-Converting Enzyme Inhibitors or Angiotensin-Receptor Blockers on SARS-CoV-2 Infection in Adults: A Living Systematic Review. Ann. Intern. Med 020 Aug 4;173(3):195–203. doi:10.7326/M20-1515. Epub 2020 May 15. PMID: 32422062; PMCID: PMC7249560.

16. Baral R, White M, Vassiliou VS. Effect of Renin-Angiotensin-Aldosterone System Inhibitors in Patients with COVID-19: a Systematic Review and Meta-analysis of 28,872 Patients. Curr. Atheroscler. Rep 2020;22(10):61. doi:10.1007/s11883-020-00880-6

17. Reynolds HR, Adhikari S, Pulgarin C, et al. Renin–Angiotensin–Aldosterone System Inhibitors and Risk of Covid-19. N. Engl. J. Med 2020;382(25):2441–48. doi:10.1056/NEJMoa2008975

18. Rentsch CT, Kidwai-Khan F, Tate JP, et al. Covid-19 Testing, Hospital Admission, and Intensive Care Among 2,026,227 United States Veterans Aged 54-75 Years. MedRxiv 2020.04.09.20059964; doi: https://doi.org/10.1101/2020.04.09.20059964

19. Mancia G, Rea F, Ludergnani M, et al. Renin–Angiotensin–Aldosterone System Blockers and the Risk of Covid-19. N. Engl. J. Med 2020;382(25):2431–40. doi:10.1056/NEJMoa2006923

20. Fosbøl EL, Butt JH, Østergaard L, et al. Association of Angiotensin-Converting Enzyme Inhibitor or Angiotensin Receptor Blocker Use With COVID-19 Diagnosis and Mortality. JAMA 2020 doi:10.1001/jama.2020.11301

21. Hippisley-Cox J, Young D, Coupland C, et al. Risk of severe COVID-19 disease with ACE inhibitors and angiotensin receptor blockers: cohort study including 8.3 million people. Heart 2020;106(19):1503–11. doi:10.1136/heartjnl-2020-317393

22. Griffith G, Morris TT, Tudball M, et al. Collider bias undermines our understanding of COVID-19 disease risk and severity. medRxiv 2020:2020.05.04.20090506. doi:10.1101/2020.05.04.20090506

23. Herbert A, Griffith G, Hemani G, et al. The spectre of Berkson’s paradox: Collider bias in Covid-19 research. Signif 2020;17(4):6–7. doi:10.1111/1740-9713.01413

24. Boëlle P-Y, Souty C, Launay T, et al. Excess cases of influenza-like illnesses synchronous with coronavirus disease (COVID-19) epidemic, France, March 2020. Euro Surveill 2020;25(14):2000326.

25. Herrett E, Gallagher AM, Bhaskaran K, et al. Data Resource Profile: Clinical Practice Research Datalink (CPRD). Int. J. Epidemiol 2015;44(3):827–36. doi:10.1093/ije/dyv098

26. Herrett E, Thomas SL, Schoonen WM, et al. Validation and validity of diagnoses in the General Practice Research Database: a systematic review. Br. J. Clin. Pharmacol 2010;69(1):4–14. doi:10.1111/j.1365-2125.2009.03537.x. PMID: 20078607; PMCID: PMC2805870

27. Clegg A, Bates C, Young J, et al. Development and validation of an electronic frailty index using routine primary care electronic health record data. Age and Ageing 2016;45(3):353–60. doi:10.1093/ageing/afw039

28. Charlson ME, Pompei P, Ales KL, et al. A new method of classifying prognostic comorbidity in longitudinal studies: development and validation. J. Clin. Epidemiol 1987;40(5):373-83. doi:https://doi.org/10.1016/0021-9681(87)90171-8

29. Khan NF, Perera R, Harper S, et al. Adaptation and validation of the Charlson Index for Read/OXMIS coded databases. BMC Fam. Pract 2010;11(1):1-7. doi:https://doi.org/10.1186/1471-2296-11-1

30. Khunti K, Singh AK, Pareek M, et al. Is ethnicity linked to incidence or outcomes of covid-19? BMJ 2020;369:m1548. doi:10.1136/bmj.m1548

31. Raisi-Estabragh Z, McCracken C, Bethell MS, et al. Greater risk of severe COVID-19 in Black, Asian and Minority Ethnic populations is not explained by cardiometabolic, socioeconomic or behavioural factors, or by 25 (OH)-vitamin D status: study of 1326 cases from the UK Biobank. J. Public Health 2020;42(3):451-60. doi:https://doi.org/10.1093/pubmed/fdaa095

32. Zhang X, Yu J, Pan LY, et al. ACEI/ARB use and risk of infection or severity or mortality of COVID- 19: A systematic review and meta-analysis. Pharmacol Res 2020;158:104927. doi:10.1016/j.phrs.2020.104927 [published Online First: 2020/05/19]

33. Chan CK, Huang YS, Liao HW, et al. Renin-angiotensin-aldosterone System Inhibitors and Risks of SARS-CoV-2 Infection: A Systematic Review and Meta-analysis. Hypertension 2020. doi:10.1161/hypertensionaha.120.15989 [published Online First: 2020/09/02]

34. Dambha-Miller H, Albasri A, Hodgson S, et al. Currently prescribed drugs in the UK that could upregulate or downregulate ACE2 in COVID-19 disease: a systematic review. BMJ open 2020;10(9):e040644. doi:10.1136/bmjopen-2020-040644 [published Online First: 2020/09/16]

35. Hasan SS, Kow CS, Hadi MA, et al. Mortality and Disease Severity Among COVID-19 Patients Receiving Renin-Angiotensin System Inhibitors: A Systematic Review and Meta-analysis. Am. J. Cardiovasc. Drugs 2020. doi:10.1007/s40256-020-00439-5 [published Online First: 2020/09/13]

36. Amat-Santos IJ, Santos-Martinez S, López-Otero D, et al. Ramipril in High-Risk Patients With COVID-19. J Am Coll Cardiol 2020;76(3):268–76. doi:10.1016/j.jacc.2020.05.040 [published Online First: 05/26]

